# Slow Gamma Suppression Correlates with Therapeutic Response to Anterior Thalamic Deep Brain Stimulation in Intractable Epilepsy

**DOI:** 10.1101/2025.08.06.25329453

**Authors:** Zachary T Sanger, Xinbing Zhang, Thomas Lisko, Hafsa Farooqi, Steffen Ventz, Sandipan Pati, Thomas Henry, David Darrow, Michael Park, Theoden I Netoff, Robert A McGovern

## Abstract

In drug-resistant epilepsy patients who undergo anterior thalamic deep brain stimulation (ANT-DBS), efficacy is assessed months after therapy initiation and clinicians have no guidance when choosing stimulation parameters due to the lack of real-time biomarkers. Here, we identified acute and chronic suppression of slow gamma oscillations (SGOs) (20–50Hz) in the ANT as a novel electrophysiological biomarker correlated with therapeutic response. Through analysis of an ongoing prospective ANT-DBS parameter optimization trial (N=11), 6/7 participants exhibiting SGOs were responders. Progressive suppression (“gamma fade”) of SGOs under chronic stimulation correlated with long-term seizure reduction in 5/6 responders. Acute stimulation in-clinic with multiple settings suppressed SGOs in 4/5 responders, challenging fixed-programming paradigms, with only one responder using the clinical gold standard parameters at the last follow-up visit. These findings establish SGO suppression as a potential multiscale biomarker for responder identification, parameter titration, and therapeutic tracking for precise, biomarker-guided intervention.

## Main

Despite advances in pharmacological and surgical therapies, epilepsy remains refractory in approximately one-third of patients (1–3), many of whom are ineligible for or have failed resective surgery (4,5). This population, disproportionately affected by uncontrolled seizures, heightened risk of sudden unexpected death in epilepsy (SUDEP), and substantial psychosocial burden, represents a persistent and growing global challenge. Neuromodulation therapies, such as vagus nerve stimulation (6–8) (VNS), responsive neurostimulation (RNS) (9–13), and deep brain stimulation (DBS) (14–16), offer a promising therapeutic alternative for individuals with drug-resistant epilepsy. However, their clinical deployment remains fundamentally constrained by a critical limitation: unlike movement disorders, where the effects of stimulation are immediately observable (e.g., tremor suppression in Parkinson’s disease), the therapeutic response to anterior thalamic (ANT) DBS unfolds gradually over weeks to months, without an accessible biomarker to guide stimulation optimization within the clinic or over time. As a result, stimulation titration is effectively blinded, relying on delayed seizure outcomes and subjective reports, which prolongs exposure to uncontrolled seizures, a period of elevated mortality and morbidity, while delaying therapeutic benefit.

This temporal disconnect introduces significant clinical uncertainty. Stimulation parameters are typically adjusted empirically, drawing on fixed protocols from pivotal trials (14), tolerability thresholds, or retrospective seizure diaries. This approach delays treatment refinement and prolongs exposure to ineffective therapy. The absence of a physiologic signal linking acute stimulation to long-term seizure outcomes has hindered progress in three essential domains: (1) rapid, data-driven stimulation titration; (2) early identification of treatment responders; and (3) development of closed-loop neuromodulation strategies tailored to individual network states. While surrogate metrics such as impedance changes (17), autonomic fluctuations (18–20), and spectral power dynamics (21–28) have shown potential for seizure detection and/or forecasting, none have provided a reliable or targetable marker of stimulation efficacy.

Here, we bridge this translational gap by identifying slow gamma oscillations (SGOs) (20–50 Hz) in the ANT, oscillatory activity measurable during routine clinical programming, whose acute suppression is observable in the clinic and chronic attenuation (‘gamma fade’) is associated with long-term seizure reduction. These findings establish SGOs as a putative biomarker of ANT-DBS efficacy, providing a rational, physiology-based framework for stimulation therapy optimization. By enabling immediate and long term feedback on therapeutic engagement, this biomarker has the potential to transform epilepsy neuromodulation from a delayed, empirically guided process into a precision therapy informed by target engagement biomarkers.

## Results

### Patient Disposition

Between 2021 and 2024, fourteen adults (≥18 years) with medically intractable focal epilepsy and clinically indicated ANT-DBS implants were prospectively enrolled in an ongoing IRB-approved clinical trial at the University of Minnesota evaluating stimulation parameter optimization. Of the 14 enrolled, 11 participants were included and 3 participants excluded from analysis due to insufficient in-clinic data and lack of seizure frequency tracking. All 11 participants had bilateral ANT-DBS leads implanted and were enrolled with Medtronic sensing enabled Percept™ systems.

Stimulation was enabled at the first neurologist follow up visit, as shown in Figure 1A, using clinically standard settings based on the SANTE pivotal trial (145 Hz, 90 us) (14). After enrollment, the participant’s stimulation amplitude was then titrated by the neurologist over follow-up visits based on the seizure frequency feedback from the participant and side effect profile. Participant demographics, seizure onset, seizure types, devices, and clinical treatment plan are shown in Table 1.

**Figure 1:**
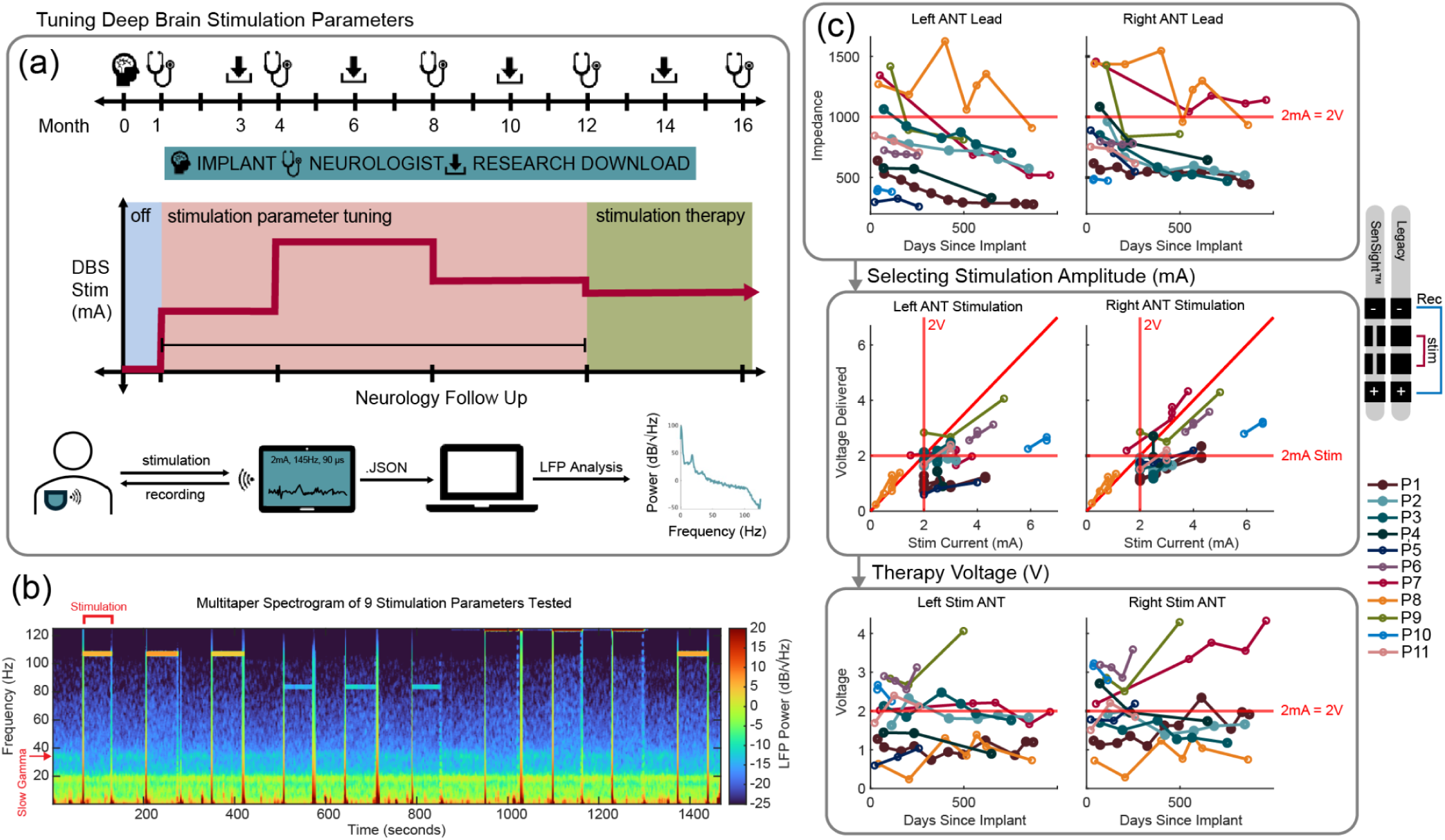
Participant Visits and ANT DBS Therapy Tuning. A) Intracranial signals were recorded at each clinical visit following implantation of their Percept™ PC device. Neurologist visits typically occur one month after implant and then repeated every 3-4 months thereafter. Data is downloaded from the Percept™ device at each of these visits storing up to 60 days of neurological signals stored at 10 minute intervals. B) Spectrogram of neural signals recorded at 250 Hz in clinic while applying different stimulation parameters. A higher LFP power is represented with a warmer color. Note the peaks around 105, 85, and 125 Hz occur during stimulation at those frequencies (or aliased from stimulation frequencies above Nyquist), while lower frequencies indicate power from neural sources. C) Left and right ANT DBS lead contacts 1 and 2 impedance across in-clinic follow up visits. The conversion between mA and volts is represented as a red line when assuming a 1kΩ electrode impedance. The trade off given the lead impedance between delivered voltage and stimulation current is shown in the second row of figures. Lastly, the delivered stimulation voltage across visits is shown for both hemispheres in all participants.

**Table 1.**
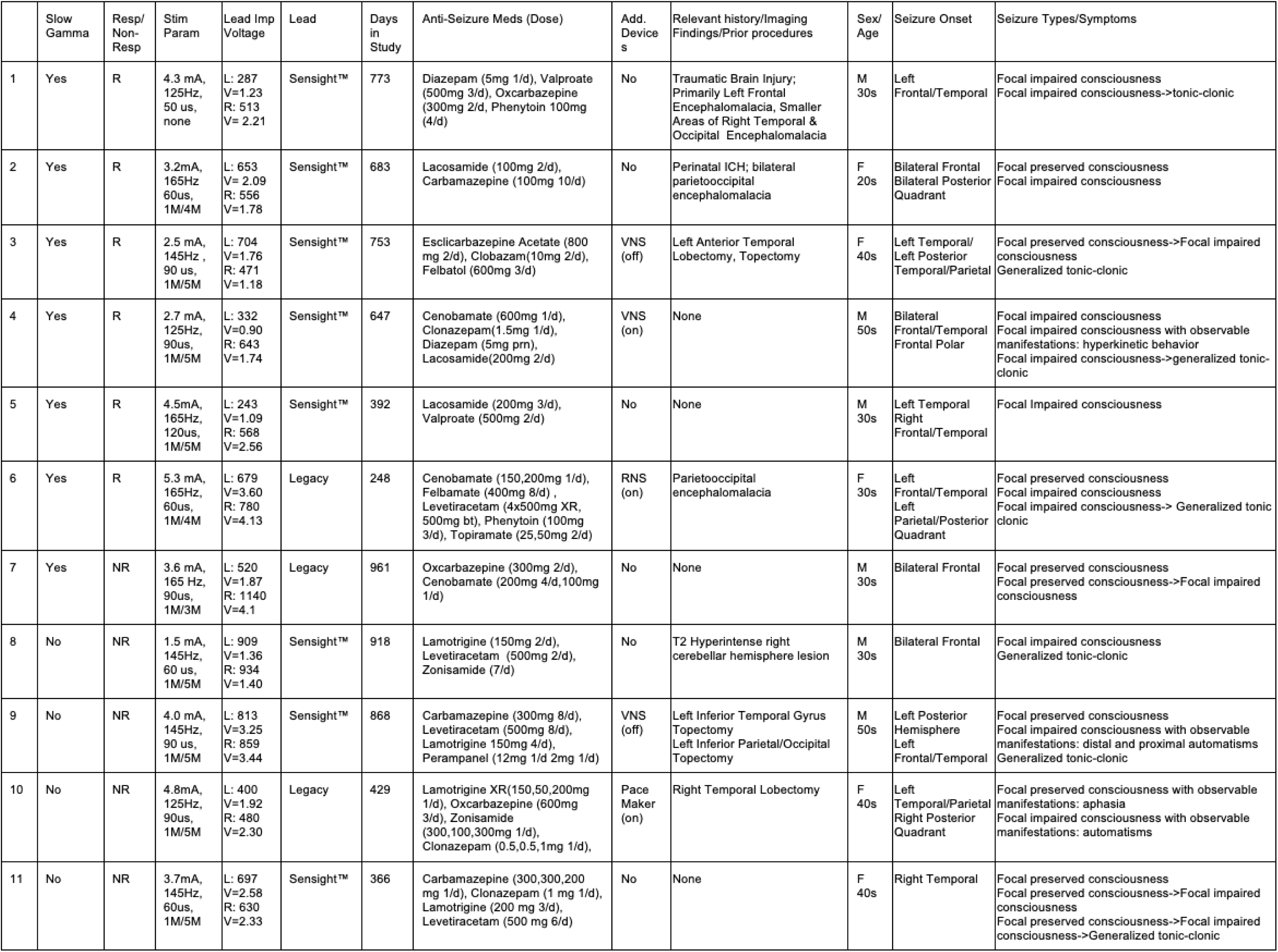
Participant Population.

At each follow-up visit, participants underwent a standardized stimulation parameter test set of 9 settings (125,145,165Hz, 60,90,120us) centered around the clinical standard SANTE trial stimulation parameter of 145Hz, 90us, as shown in Figure 1A&B. To estimate the changes in participants’ broadband LFP response to different stimulation parameters, a gaussian process regression fit was used to estimate a response surface drawing from the mean LFP response under the initially tested settings. Using the response surface, further testing was conducted around the stimulation settings that resulted in the greatest suppression of neural activity for each participant.

Impedance in each ANT hemisphere (lead ring 1 and 2 to allow for sensing 0-3) was recorded and plotted in Figure 1C. We observed the majority of participants with ANT lead implants exhibited impedances lower than 1kΩ, with many implants around 500Ω. While electrical stimulation therapies utilize constant current stimulation for the safety benefits, therapy efficacy may be dependent on the delivered electrical field (volts/meter) and its distribution through the local neural tissue as opposed to the current amplitude defined (29). This trade off in each participant between the current programmed and delivered voltage is shown based on the impedance measurement extracted by the Percept™ device as shown in Figure 1C. As a result, many participants were programmed with amplitudes ranging from 4-6 mA to then receive approximately 2-3 volts of delivered electrical stimulation.

### Local Field Potentials and Seizure Frequency

While testing different stimulation parameters across follow-up visits, we recorded bilateral ANT LFPs during stimulation on and off conditions. LFP activity was sensed in each participant through the 0 and 3 ring contacts of the lead. Lead placement is shown in Figure 2A within a standardized Morel atlas using LeadDBS. The leads were then colored red (left) or green (right) if a SGO (20-50Hz) peak was observed in the LFP multitaper power spectral density (PSD) estimate.

**Figure 2:**
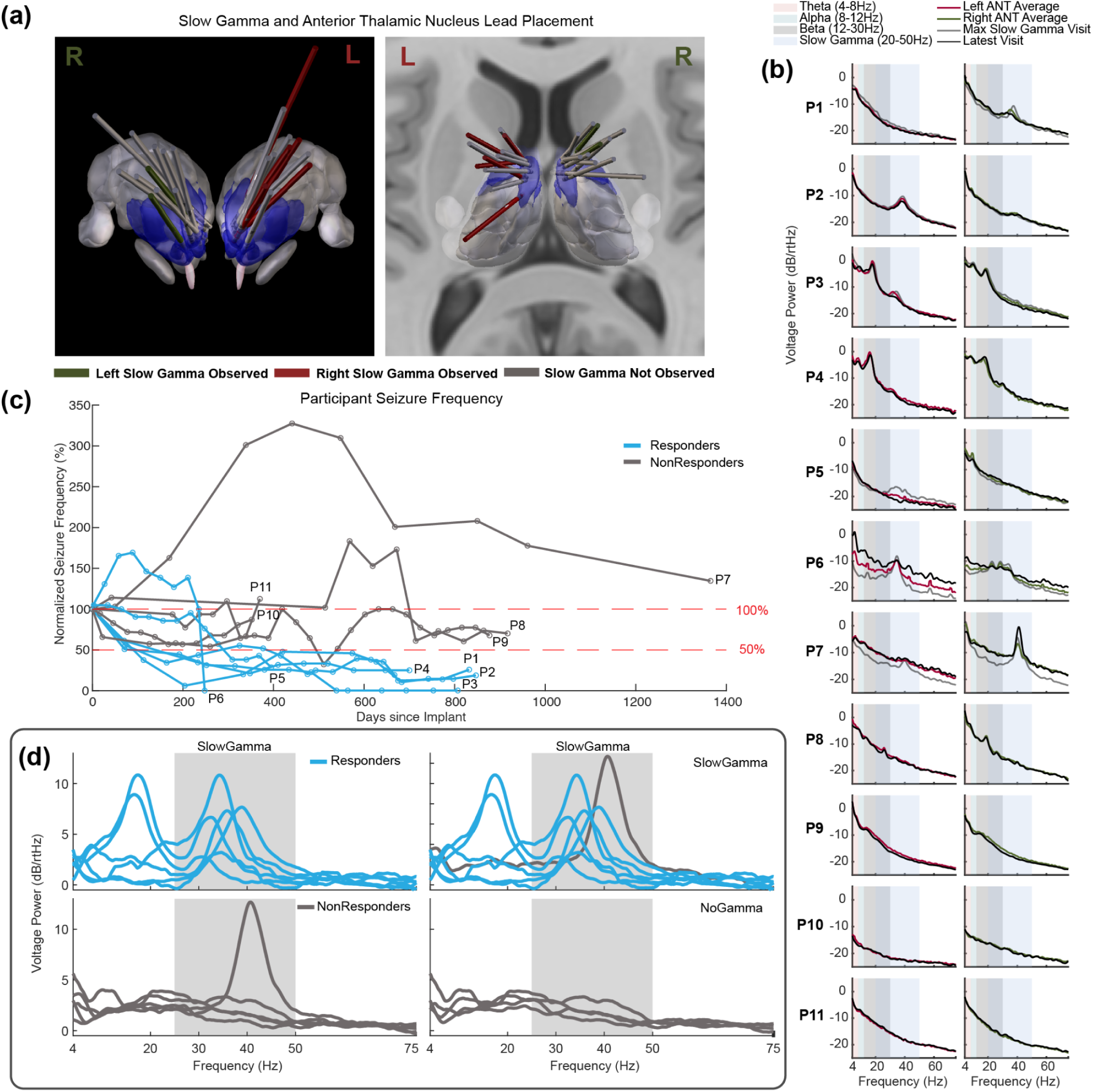
Observed Neural Activity in Responders and Non-Responders to ANT DBS. A) Coronal and axial views of all participants’ bilateral lead placements plotted using a standardized atlas with LeadDBS. Red (left) and green (right) leads demonstrate where SGOs were observed. Blue shaded tissue represents the ANT. B) PSD responses without stimulation across all participants with the neural Berger bands are shown for biomarker identification. For each participant, the average and latest PSD is shown with an additional max SGO PSD in participants where SGOs were observed. C) Seizure reduction normalized by the pre-DBS monthly seizure frequency for all participants. Blue represents responders and gray for non-responders. D) Participants are grouped by their max 1/*f* baseline SGO detrended PSDs based on responder/nonresponder or SGO/no SGO categorization. The colors represent each participant’s responder status based on 2C.

Across the 11 participants, we observed a SGO peak in 7 participants (P1-P7) in the baseline PSD response with stimulation off as shown in Figure 2B. This SGO peak was predominantly in one hemisphere, not consistently on a particular side, with varying peak frequencies. The left and right ANT PSD is shown with the max SGO visit, average PSD across visits, and most recent visit PSD for each participant. To better understand the utility of this SGO response during stimulation, participant seizure frequency was recorded through seizure diaries at every research visit and is shown in Figure 2C. Participants were deemed a responder to ANT-DBS if they achieved a seizure reduction greater than 50% by their last follow-up visit. To reduce reporting variability and seizure relapsing, a 3 sample moving average was used to extract seizure frequency from the raw reported data. Some non-responders had a short period of 50% reduction, however, quickly relapsed back to their pre-DBS seizure frequency.

A Fisher exact test was used to understand the association between responders and those with an observable baseline SGO peak. There was a statistically significant association between SGOs and responder status (p=0.015).

Participants were sorted into groups based on responder status and whether SGOs were observed as shown in Figure 2D. Of the 7 participants with observable SGOs, 6 were responders. Notably, the only non-responder in this group was the one participant in whom we could not modulate SGO activity with DBS. All of the participants without SGOs were non-responders.

### DBS Modulation of ANT Slow Gamma Oscillations (SGOs)

While observation of an SGO signal in the ANT may be a baseline signal seen in responders, we were interested in ANT-DBS’s ability to modulate this neural signal. In Figure 3A, the mean SGO LFP PSD response is shown across the frequency versus pulse width response surface. Individualized research-optimized stimulation parameters based on the SGO LFP are shown on each surface as a red circle. Participants P1-P3 demonstrated larger baseline SGOs and SGO suppression, leading to higher dB differences between stimulation parameters.

**Figure 3:**
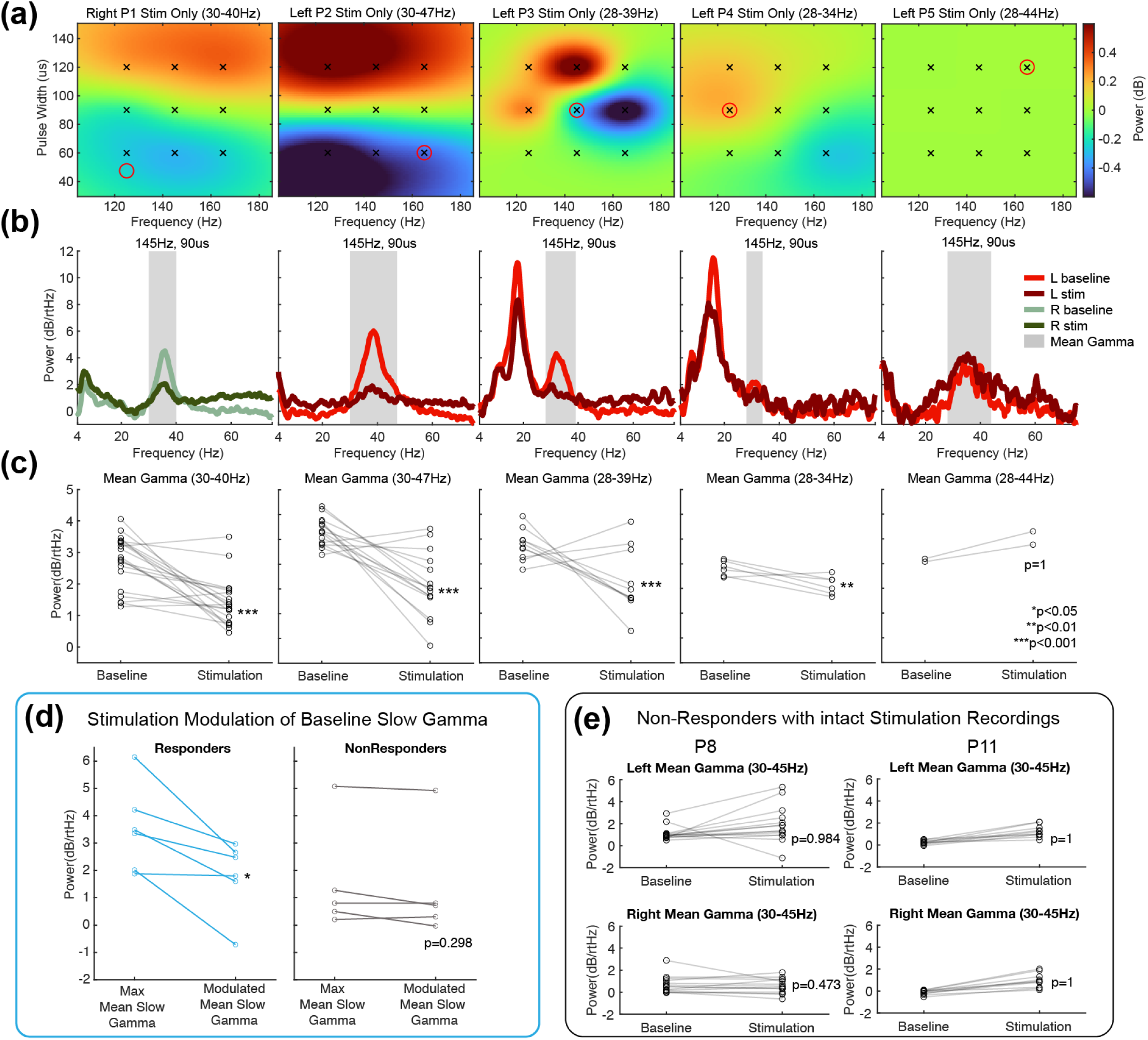
Slow Gamma Modulation in the Clinic. A) In all responder participants without stimulation recording artifacts, average gaussian process regression surfaces across visits for each participant where SGOs were observed are shown. Each SGO frequency range is shown in the title and also shown as a shaded region in 3B. The black x’s represent the stimulation parameter test set and the red circle indicates their latest setting. Cool colors demonstrate a larger reduction in SGOs as compared to the other stimulation parameters. B) Average baseline and stimulation 1/*f* detrended PSD under 145Hz, 90us across participants in 3A. C) Mean SGO response in each baseline and stimulation test pair across visits with the associated Wilcoxon rank sum one-sided p-value for SGO suppression during stimulation. D) Modulation of SGOs shown across all participants organized by responders and non-responders. A Wilcoxon rank sum comparison between the max observed SGO and most recently recorded SGO LFP response in both groups to show change in SGO across visits under stimulation. E) Bilateral SGO changes in two non-responders during ANT-DBS under the clinical setting. Non-significant p-values determined through a Wilcoxon sum rank test.

The standard ANT DBS clinical setting, 145Hz, 90us, is shown in Figure 3B across these participants to demonstrate the average PSD response observed across all visits during standard clinical stimulation. Figure 3C demonstrates the change between baseline and stimulation for each stimulation setting test across all the visits shown in Figure 3B. A Wilcoxon rank sum test demonstrated a single tail statistically significant reduction in SGOs during stimulation (P1: p=5.8E-5, P2: p=3.4E-5, P3: p=7E-3, P4: p=4E-3, P5: p=1). Participant P6 and P7 stimulation modulation could not be determined due to multiple artifacts throughout all stimulation recordings. All P6 and P7 PSDs during stimulation had a roughly 5dB greater background noise level than during baseline, resulting in the noise floor completely covering an observed SGO signal during baseline.

The chronic effects of stimulation on SGOs in all participants, not just participants where stimulation recordings were not corrupted, are shown in Figure 3D. Statistically significant suppression of SGOs were observed in responders when comparing the visit with max mean SGOs compared to their current mean SGOs. Lastly, in participants with non-corrupted stimulation LFP responses that were not responders, no acute statistically significant SGO suppression was observed as shown in Figure 3E.

### “Gamma Fade”: ANT DBS Suppression of SGOs Over Time Correlates with Seizure Reduction

Not only can SGO reduction be observed acutely within the clinic, but a steady reduction in baseline SGOs was observed across visits. During in-clinic follow-up, temporarily disabling stimulation allowed for observation of baseline SGO activity changes over time under chronic ANT DBS therapy. We have named this reduction “gamma fade”, as the impact of stimulation over multiple visits suppressed the SGO activity observed over time. Across visits within responders, baseline SGO power slowly decreased and SGOs observed during stimulation were consistently suppressed as shown in Figure 4A.

**Figure 4:**
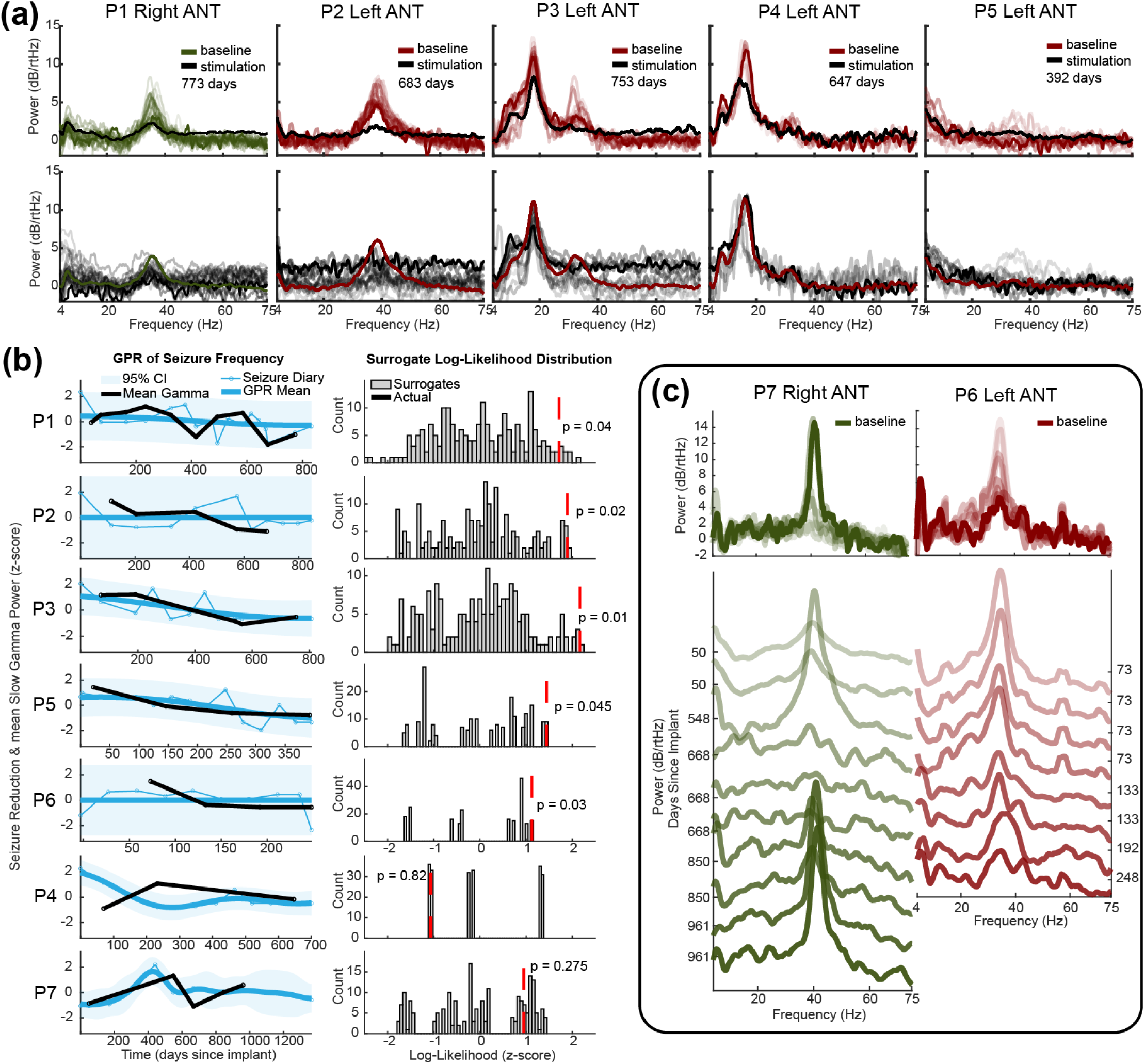
Baseline Gamma Fade across Visits. A) Responder participants with Sensight™ leads and an SGO peak with the change in SGOs during stimulation and baseline across visits. A more transparent detrended PSD (Right (green) and Left (red) represents an earlier clinical visit. Baseline is shown with an average stimulation PSD over visits (top) or stimulation is shown with an average baseline PSD (bottom). In participants P2 and P3, a rise in the noise floor is observed at higher frequencies. B) Seizure reduction normalized by the pre-DBS seizure frequency correlated with 5/7 SGO participants is shown. Seizure diary reported seizure reduction is shown in blue correlated in time with the observed mean visit baseline SGO power in black. For each participant, a histogram of the log likelihood of predicting the mean SGO power from the seizure reduction model is shown. The log likelihood of slow-gamma time shuffled surrogates are compared to the log likelihood of the actual data (shown as a red dotted line). C) Legacy lead participants, one responder (P6) and one non-responder (P7) baseline detrended PSD across visits. Stimulation is not shown due to artifact corruption during stimulation.

To investigate the connection between the baseline SGO fade and the patient’s improvement with DBS, we assessed the seizure reduction response’s ability to predict the mean SGO amplitude in the clinic across visits. Five of six responders showed statistically significant correlation of seizure frequency with mean SGOs (Figure 4B). Four of five non-responders demonstrated no SGO peaks and therefore this relationship could not be assessed. The one non-responder who did have a SGO peak (P7, Figure 2B,C,E) that was unable to be modulated with stimulation showed no significant relationship between seizure frequency and mean SGOs.

Lastly, participants P6 and P7 are participants with legacy leads and exhibited multiple artifacts during stimulation, rendering acute stimulation effects unobservable. With stimulation off, their baseline activity can be observed (Figure 4C) over multiple visits. Participant P7 is the only participant in our dataset that exhibited gamma that has gotten worse alongside a fluctuating seizure frequency and more severe seizures. Importantly, stimulation did not appear to affect SGO power amplitude throughout their course. Participant P6 has demonstrated consistent SGO reduction since receiving their Percept™ system, allowing us to record in-clinic baseline LFP signals.

## Discussion

Our discovery of ANT SGOs as a biomarker of DBS efficacy redefines therapeutic paradigms for drug-resistant epilepsy by resolving a fundamental disconnect: the inability to assess stimulation effects in real time. We demonstrate that SGOs operate across three clinically actionable timescales: as a potential predictor prior to implant (85.7% of participants with SGOs early in therapy delivery were responders), a dose-titration signal (acutely suppressed by effective parameters), and a longitudinal tracker (exhibiting progressive "gamma fade" with therapeutic benefit). This multiscale utility addresses the critical limitation of current practice, where clinicians optimize DBS blindly while patients remain at elevated risk of seizures during months of titration.

The strong association between baseline SGOs and response (p=0.015) suggests thalamic SGOs may identify patients with ANT-connected epileptogenic networks. Notably, the biomarker’s value lies not merely in detection but in modifiability. The non-responder with unmodifiable increasing SGOs over time (P7) underscores that this oscillation may be pathologic and must be disrupted to provide therapeutic benefit. This aligns with emerging DBS principles where biomarker responsiveness, not just presence, determines clinical utility. While SGOs were suppressed with the clinical and multiple research parameters in the clinic, chronic suppression of SGOs was demonstrated under patient-specific parameters in 5/6 responders, challenging the fixed-parameter approach of historical trials. This also lays the foundation for establishing patient-specific stimulation paradigms for future trials.

While our findings demonstrate a correlation between participants’ SGO peak modulation by ANT DBS and response rate, the origin of this slow-gamma oscillation could not be identified in this study because recordings were limited to the ANT. Furthermore, it is not known how it correlates with seizure severity, seizure types, seizure onset or the mechanism by which it fades over time. The "gamma fade" phenomenon (progressive baseline reduction correlating with seizure improvement) suggests ANT-DBS induces enduring network reorganization. No prior studies have examined this phenomenon in humans, but one pilot study in a single ANT-DBS responder (81% seizure reduction) demonstrated a strong slow-gamma peak and fade over time, consistent with our findings (30). Though not specifically noted in the manuscript, this does independently illustrate these results.

The ANT is well connected within the hippocampal circuit of Papez with bidirectional connections to the amygdala and mesial frontal cortical structures, and receives inputs through the mammillothalamic tracts, thalamocortical radiations, and the fornix (31,32). Functionally, the ANT is believed to facilitate communication through theta synchrony between mesial temporal and neocortical structures (32–35). While it is a possibility that this SGO occurs locally, current ANT DBS mechanisms hypothesize that ANT DBS inhibits incoming pathological activity and restores normal physiological outputs from the ANT (36,37). These SGOs may not originate in the ANT, but arrive through input connections or are generated through reciprocal connections with another area.

Slow gamma rhythms are seen in the hippocampus and arise through connections between the CA1,CA3, and dentate gyrus (38–41). Given that the hippocampus connects to the ANT through the fornix and mammillothalamic tracts, this could be one possible origin of the slow-gamma oscillations in ANT. Further supporting this, one study has shown that anterior dorsal placement of ANT-DBS leads resulted in outcomes with the highest seizure reduction (42) and this area receives inputs from the mammillothalamic tracts (43). Proper placement has been verified post implant in ANT-DBS participants by utilizing hippocampal evoked potentials during ANT stimulation, demonstrating that target placement contains strong connections from the hippocampus (44). Additionally, case studies have shown that patients with strong hippocampal evoked potentials during ANT-DBS have been correlated with good seizure control (45). Combined, these results support the hypothesis that SGOs originate within the circuit of Papez and can lead to successful ANT-DBS outcomes. If these oscillations arrive from the hippocampus, then hippocampal decoupling observed in responders with other neuromodulatory therapies may explain the improvement we observe.

However, the anterior nucleus of the thalamus is bidirectionally connected to the neocortex through thalamocortical radiations and connections with the cingulate cortex (46). The cingulate cortex, a region with bidirectional connections to the ANT within the circuit of Papez, has been shown to be well connected to the frontal, temporal, and parietal cortices (47). A pediatric epilepsy study found a correlation between frontotemporal lobe epilepsy with impaired executive function and damaged, less developed thalamocortical pathways (48). Another study in frontal lobe epilepsy participants found that after 3 years, 14/16 participants who underwent anterior thalamic radiation disconnection as part of their frontal lobe resections were seizure-free, whereas only 11/31 were seizure free in patients who had those tracts preserved (49). During working memory tasks, SGOs have also been observed in the medial prefrontal cortex coupled to theta oscillations within the hippocampus (50). Therefore, not only do thalamocortical radiations play a role in frontotemporal lobe epilepsy, but cortical structures associated with executive function tasks may generate SGOs.

SGO observation raises the provocative question of whether chronic SGO suppression disrupts pathologic thalamocortical synchrony. The correlation between fade kinetics and seizure reduction timing hints at plastic changes underlying therapeutic effects, though causal studies are needed. Mechanistically, SGOs may reflect aberrant thalamic gating of cortico-limbic traffic; its suppression could normalize hyperexcitable circuits.

Translational opportunities are immediate: sensing-enabled devices (e.g., Percept™) already capture this signal, enabling prospective trials comparing SGO-guided versus empiric titrated stimulation parameters. Future work should investigate whether pre-implant network mapping (e.g., tractography) and/or seizure network localization through stereo-electroencephalography (sEEG) can predict "gamma positive" phenotypes, optimizing patient selection for ANT DBS. Pre-implant SGO signal localization may also foster investigation into new stimulation targets interconnected with the ANT.

There are limitations to this study. First, although the relationship between SGO suppression and seizure reduction was robust across participants, causal inference is limited by the observational design. Whether suppression of SGOs is necessary and sufficient for therapeutic effect remains to be tested in a large sample size. Second, artifact contamination during stimulation limited analysis in two participants, emphasizing the need for improved artifact mitigation strategies in chronic sensing systems. Artifact contamination in discontinued, legacy leads will diminish with next-generation directional electrodes and artifact-suppression algorithms. Third, while our findings are promising, the sample size is modest, and larger multicenter studies will be necessary to validate the generalizability of SGOs as a biomarker. Additionally, the clinical utility of this biomarker in real-time closed-loop paradigms has yet to be explored. In one participant, we observed SGOs were maximally observable during the day in a participant with primarily nocturnal seizures (51). Others have demonstrated the positive effects on ictal activity through stimulation during interictal states (52). Further investigation of SGO suppression adaptively during interictal states could preserve efficacy while reducing side effects like ANT-DBS sleep disruption (53).

This work establishes SGOs as both a biomarker and a fundamental axis of network pathophysiology, bridging acute ANT-DBS effects to chronic seizure control. The convergence of adaptive stimulation with SGO tracking can push this field into a new era of dynamic neuromodulation, where therapy adapts not merely to disease states but to individual network dynamics in real time. Crucially, this approach shifts the paradigm from reactive seizure suppression to proactive circuit modulation, tuning stimulation to electrophysiological states of therapeutic readiness rather than clinical events. SGOs thus provide the first evidence-based roadmap for closed-loop DBS in epilepsy, offering a measurable substrate to accelerate optimization, personalize parameters, and ultimately mitigate the prolonged mortality risk inherent to traditional trial-and-error programming.

## Methods

### In Clinic Visit

All participant data (N=11) shown has been collected as part of an ongoing University of Minnesota clinical trial (#NCT05493722) investigating the efficacy of personalized DBS stimulation parameters to reduce the broadband power within the Anterior Nucleus of the Thalamus (ANT). Within this trial, a large LFP dataset collected from many refractory epilepsy participants has provided the opportunity to retrospectively analyze any biomarkers that may be present and how DBS neuromodulation may impact potential biomarkers. The analysis shown here is exploratory and not the original intent of the trial, however, this retrospective analysis may aid in future clinical trials to specifically investigate this analysis.

Participants have all received Medtronic Percept™ Implantable Pulse Generators (IPG) for its ability to simultaneously stimulate and record. Some participants were implanted with Medtronic DBS Legacy leads (N=2, 3389) and others received the latest Medtronic Sensight™ DBS leads (N=9, B33005).

The LFP data shown here is composed of immediate baseline recordings and stimulation recordings of 9 different settings tested around the SANTE trial (14) clinical setting (145Hz, 90us). The 9 setting 3x3 setting grid is limited by the Medtronic Percept™ BrainSense™ artifact reducing stimulation parameter options: 125Hz, 145Hz, or 165Hz and 60us, 90us, or 120 us pulse width. Stimulation amplitude was titrated clinically accordingly by the physician across follow up visits. Electrode impedance measurements were taken at each follow up and participant seizure diary was documented by the research team.

For each setting, 1 minute of baseline activity was recorded prior to each setting with 1 minute of stimulation. Participants were awake, attentive, and interacting with the staff throughout data collection. Stimulation amplitude, frequency, and pulse width were adjusted across different settings while preserving the total electrical energy delivered (TEED). During in-clinic recordings, stimulation cycling and soft start were disabled because the participants could tolerate this change and it reduced potential artifacts observable in the LFP recordings.

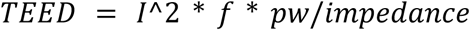

Acute effects of stimulation could not be studied in participants with Legacy leads due to artifacts that caused broadband shifts in the noise floor. For these participants, results in figures 2, 3 and 4 only show changes with stimulation off.

### LFP Pre-processing and Signal Analysis

For improved neural oscillation visualization and comparison of stimulation parameters, we developed a pre-processing and analysis pipeline based on pre-existing tools in the neuromodulation field. Epilepsy participants’ neural activity in the time domain, as compared to other neural conditions, can manifest differently depending on the individual’s seizure activity landscape. Additionally, Medtronic legacy leads were not designed to be utilized for neural recording and it has been shown that these leads can be susceptible to fluid ingress (54–56). We have observed ECG artifacts in participants with ANT DBS Sensight™ leads (57). So, when QRS cardiac complexes were visually perceivable in the raw LFP recordings, unilateral or bilateral cardiac artifacts were removed using a template subtraction method in the time domain.

The PSD of the time domain LFP data was then estimated using a Short Time Fourier Transform frequency transform approach using multiple orthogonal windowing functions, often referred to as the multitaper transform (57). This approach provided lower PSD variance while maintaining frequency main lobe resolution at the cost of the phase data. Low frequency artifact such as stimulation on/off switching transients, ictal discharges, or movement artifact can be difficult to characterize, so these artifacts were removed in the frequency domain by rejecting windows across the short time fourier transform response before averaging across times to determine the LFP recording PSD.

Once the filtered PSD was generated for each stimulation trial with an associated prior baseline, the PSD was detrended into the periodic component using a least squares regression fit approach with the Fitting Oscillations and One Over F fitting (FOOOF) (58) logarithmic function to estimate the *1/f* aperiodic component (57).

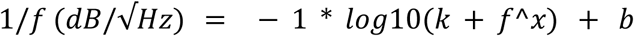

Because the aperiodic component is believed to be the background activity of that brain region, the aperiodic fit of the baseline recording was used to detrend both the baseline and stimulation responses for each setting in the event stimulation had an effect on the 1/f shape.

### Statistical Analysis and Gaussian Process Regression Estimation

To assess the association between being a responder and having an SGO baseline response, a Fisher exact test was used comparing responders/nonresponders and SGO/no SGO participants in the population.

An LFP response surface of the participant’s frequency and pulse width parameter space was estimated using a gaussian process regression fit of the ANT LFP response to 9 initial stimulation parameter configurations. Each participant’s SGOs were band-passed based on visually determining when the SGO peak deviated from the 1/f aperiodic baseline PSD response detrended by the preceding baseline 1/f fit. A gaussian process regression was generated using a matern 3/2 kernel with a length scale of 2.5 and sigma of 0.1. Over time, SGOs were modulated by stimulation resulting in a decreased signal to modulate with stimulation. To assess the average relative differences between stimulation parameters across visits to modulate SGOs, visits where minimal SGO modulation was observed later in follow up were omitted. To demonstrate differences across settings, the response surface was normalized by the mean response of the 9 measured settings. If a stimulation parameter other than 145Hz, 90us exhibited a greater reduction in SGOs, that research optimized setting was tested by the participant at home.

To compare the effect of ANT DBS on SGOs, the mean SGO response within participants during baseline and stimulation was compared across visits using a single tail Wilcoxon rank sum test.

Due to sparse sampling, the seizure reduction correlation with mean SGO power suppression was calculated using a gaussian process regression model of the seizure reduction to obtain a seizure response over time. Some participants tracked their seizure frequency more frequently than they visited their neurologist. Using this model with both seizure reduction and mean SGO power z-scored, if the data was correlated, the model should effectively predict mean SGO power at a specific time during follow up. We then compared these results with time shuffled surrogates of mean SGO power to compare against mean SGO power occurring over follow up.

## Acknowledgements

Thank you to the participants and their neurologists for their contributions and participation within the study. Thank you to Scott Stanslaski for technical assistance when using the Medtronic Percept™ system within our ongoing clinical trial. Figures and technical content has been adapted or is drawn upon from Zachary Sanger’s thesis dissertation (59).

## Data Availability Statement

Data can be made available upon reasonable request to the corresponding author.

## Supplemental Figures

**Supplemental Figure 1:**
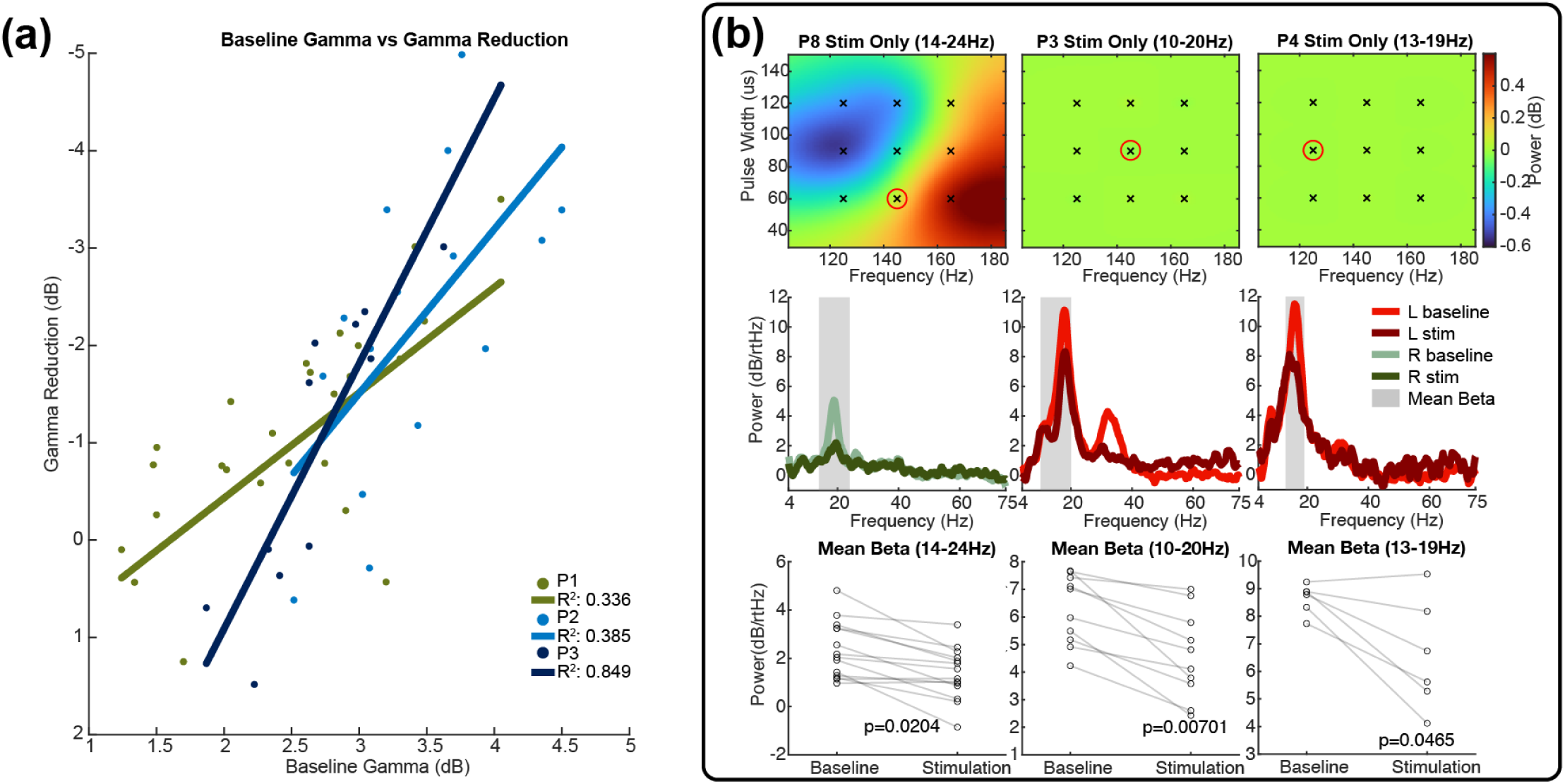
ANT DBS LFP Modulation. A) Linear regression of responder participants 1, 2, and 3 demonstrated large SGO reduction across multiple visits to show the relationship between baseline SGOs and the amount of SGO reduction observed. B) In participants 8 (NR), 3 (R), and 4 (R), statistically significant beta suppression is shown. Relative beta suppression across the 9 tested settings is shown with the 145Hz, 90us baseline and stimulation PSD shown with associated Wilcoxon rank sum p-values.

## References

1. Kwan P, Schachter SC, Brodie MJ. Drug-Resistant Epilepsy. N Engl J Med. 2011 Sep 8;365(10):919–26.

2. Dalic L, and Cook MJ. Managing drug-resistant epilepsy: challenges and solutions. Neuropsychiatr Dis Treat. 2016 Oct 12;12:2605–16.

3. Sheng J, Liu S, Qin H, Li B, Zhang X. Drug-Resistant Epilepsy and Surgery. Curr Neuropharmacol. 2018 Jan 1;16(1):17–28.

4. Wiebe S, Blume WT, Girvin JP, Eliasziw M, Effectiveness and Efficiency of Surgery for Temporal Lobe Epilepsy Study Group. A randomized, controlled trial of surgery for temporal-lobe epilepsy. N Engl J Med. 2001 Aug 2;345(5):311–8.

5. Engel J, McDermott MP, Wiebe S, Langfitt JT, Stern JM, Dewar S, et al. Early surgical therapy for drug-resistant temporal lobe epilepsy: a randomized trial. JAMA. 2012 Mar 7;307(9):922–30.

6. Panebianco M, Rigby A, Weston J, Marson AG. Vagus nerve stimulation for partial seizures. Cochrane Database Syst Rev. 2015 Apr 3;2015(4):CD002896.

7. Englot DJ, Chang EF, Auguste KI. Vagus nerve stimulation for epilepsy: a meta-analysis of efficacy and predictors of response. 2011 Dec 1 [cited 2025 Apr 15]; Available from: https://thejns-org.ezp2.lib.umn.edu/view/journals/j-neurosurg/115/6/article-p1248.xml

8. Toffa DH, Touma L, El Meskine T, Bouthillier A, Nguyen DK. Learnings from 30 years of reported efficacy and safety of vagus nerve stimulation (VNS) for epilepsy treatment: A critical review. Seizure. 2020 Dec;83:104–23.

9. Thomas GP, Jobst BC. Critical review of the responsive neurostimulator system for epilepsy. Med Devices Auckl NZ. 2015;8:405.

10. Nair DR, Laxer KD, Weber PB, Murro AM, Park YD, Barkley GL, et al. Nine-year prospective efficacy and safety of brain-responsive neurostimulation for focal epilepsy. Neurology. 2020 Sep 1;95(9):e1244–56.

11. Rønborg SN, Esteller R, Tcheng TK, Greene DA, Morrell MJ, Wesenberg Kjaer T, et al. Acute effects of brain-responsive neurostimulation in drug-resistant partial onset epilepsy. Clin Neurophysiol. 2021 Jun 1;132(6):1209–20.

12. Heck CN, King-Stephens D, Massey AD, Nair DR, Jobst BC, Barkley GL, et al. Two-year seizure reduction in adults with medically intractable partial onset epilepsy treated with responsive neurostimulation: final results of the RNS System Pivotal trial. Epilepsia. 2014 Mar;55(3):432–41.

13. Razavi B, Rao VR, Lin C, Bujarski KA, Patra SE, Burdette DE, et al. Real-world experience with direct brain-responsive neurostimulation for focal onset seizures. Epilepsia. 2020 Aug;61(8):1749–57.

14. Fisher R, Salanova V, Witt T, Worth R, Henry T, Gross R, et al. Electrical stimulation of the anterior nucleus of thalamus for treatment of refractory epilepsy. Epilepsia. 2010;51(5):899–908.

15. Salanova V, Witt T, Worth R, Henry TR, Gross RE, Nazzaro JM, et al. Long-term efficacy and safety of thalamic stimulation for drug-resistant partial epilepsy. Neurology. 2015 Mar 10;84(10):1017–25.

16. Salanova V, Sperling MR, Gross RE, Irwin CP, Vollhaber JA, Giftakis JE, et al. The SANTÉ study at 10 years of follow-up: Effectiveness, safety, and sudden unexpected death in epilepsy. Epilepsia. 2021;62(6):1306–17.

17. Mivalt F, Kremen V, Sladky V, Cui J, Gregg NM, Balzekas I, et al. Impedance Rhythms in Human Limbic System. J Neurosci Off J Soc Neurosci. 2023 Sep 27;43(39):6653–66.

18. Karoly PJ, Stirling RE, Freestone DR, Nurse ES, Maturana MI, Halliday AJ, et al. Multiday cycles of heart rate are associated with seizure likelihood: An observational cohort study. eBioMedicine. 2021 Oct 1;72:103619.

19. Brinkmann B, Nurse E, Nasseri M, Viana PF, Karoly P, Attia TP, et al. Seizure forecasting and detection with wearable devices and subcutaneous EEG - A Practical Seizure Gauge (N2.004). Neurology. 2022 May 3;98(18_supplement):528.

20. Kremen V, Sladky V, Mivalt F, Gregg NM, Brinkmann BH, Balzekas I, et al. Modulating limbic circuits in temporal lobe epilepsy: impacts on seizures, memory, mood and sleep. Brain Commun. 2025 Apr 1;7(2):fcaf106.

21. Langdon-Down M, Russell Brain W. TIME OF DAY IN RELATION TO CONVULSIONS IN EPILEPSY. The Lancet. 1929 May 18;213(5516):1029–32.

22. Griffiths GwenvronM, Fox JT. RHYTHM IN EPILEPSY. The Lancet. 1938 Aug 20;232(5999):409–16.

23. Baud MO, Kleen JK, Mirro EA, Andrechak JC, King-Stephens D, Chang EF, et al. Multi-day rhythms modulate seizure risk in epilepsy. Nat Commun. 2018 Jan 8;9(1):88.

24. Karoly PJ, Rao VR, Gregg NM, Worrell GA, Bernard C, Cook MJ, et al. Cycles in epilepsy. Nat Rev Neurol. 2021 May;17(5):267–84.

25. Kreitlow BL, Li W, Buchanan GF. Chronobiology of epilepsy and sudden unexpected death in epilepsy. Front Neurosci [Internet]. 2022 [cited 2023 May 5];16. Available from: https://www.frontiersin.org/articles/10.3389/fnins.2022.936104

26. Gregg NM, Pal Attia T, Nasseri M, Joseph B, Karoly P, Cui J, et al. Seizure occurrence is linked to multiday cycles in diverse physiological signals. Epilepsia. 2023;64(6):1627–39.

27. Satzer D, Kaye LC, Ojemann SG, Kramer DR, Thompson JA. Aperiodic activity as a biomarker of seizures and neuromodulation. Brain Stimulat. 2025 May 1;18(3):738–44.

28. Yang AI, Raghu ALB, Isbaine F, Alwaki A, Gross RE. Sensing with deep brain stimulation device in epilepsy: Aperiodic changes in thalamic local field potential during seizures. Epilepsia. 2023;64(11):3025–35.

29. Lempka SF, Johnson MD, Miocinovic S, Vitek JL, McIntyre CC. Current-controlled deep brain stimulation reduces in vivo voltage fluctuations observed during voltage-controlled stimulation. Clin Neurophysiol Off J Int Fed Clin Neurophysiol. 2010 Dec;121(12):2128–33.

30. Satzer D, Wu S, Henry J, Doll E, Issa NP, Warnke PC. Ambulatory Local Field Potential Recordings from the Thalamus in Epilepsy: A Feasibility Study. Stereotact Funct Neurosurg. 2023 May 22;101(3):195–206.

31. Majtanik M, Gielen F, Coenen VA, Lehtimäki K, Mai JK. Structural connectivity of the ANT region based on human ex-vivo and HCP data. Relevance for DBS in ANT for epilepsy. NeuroImage. 2022 Nov 15;262:119551.

32. Warsi NM, Yan H, Suresh H, Wong SM, Arski ON, Gorodetsky C, et al. The anterior and centromedian thalamus: Anatomy, function, and dysfunction in epilepsy. Epilepsy Res. 2022 May;182:106913.

33. Sweeney-Reed CM, Zaehle T, Voges J, Schmitt FC, Buentjen L, Borchardt V, et al. Anterior Thalamic High Frequency Band Activity Is Coupled with Theta Oscillations at Rest. Front Hum Neurosci. 2017 Jul 20;11:358.

34. Johnson EL, Adams JN, Solbakk AK, Endestad T, Larsson PG, Ivanovic J, et al. Dynamic frontotemporal systems process space and time in working memory. PLOS Biol. 2018 Mar 30;16(3):e2004274.

35. Sweeney-Reed CM, Buentjen L, Voges J, Schmitt FC, Zaehle T, Kam JWY, et al. The role of the anterior nuclei of the thalamus in human memory processing. Neurosci Biobehav Rev. 2021 Jul;126:146–58.

36. Bouwens van der Vlis TAM, Schijns OEMG, Schaper FLWVJ, Hoogland G, Kubben P, Wagner L, et al. Deep brain stimulation of the anterior nucleus of the thalamus for drug-resistant epilepsy. Neurosurg Rev. 2019;42(2):287–96.

37. McIntyre CC, Grill WM, Sherman DL, Thakor NV. Cellular effects of deep brain stimulation: model-based analysis of activation and inhibition. J Neurophysiol. 2004 Apr;91(4):1457–69.

38. Carr MF, Karlsson MP, Frank LM. Transient Slow Gamma Synchrony Underlies Hippocampal Memory Replay. Neuron. 2012 Aug 23;75(4):700–13.

39. Hsiao YT, Zheng C, Colgin LL. Slow gamma rhythms in CA3 are entrained by slow gamma activity in the dentate gyrus. J Neurophysiol. 2016 Dec 1;116(6):2594–603.

40. Maglóczky Z, Freund TF. Impaired and repaired inhibitory circuits in the epileptic human hippocampus. Trends Neurosci. 2005 Jun 1;28(6):334–40.

41. Freund TF, Katona I. Perisomatic Inhibition. Neuron. 2007 Oct 4;56(1):33–42.

42. Gross RE, Fisher RS, Sperling MR, Giftakis JE, Stypulkowski PH. Analysis of Deep Brain Stimulation Lead Targeting in the Stimulation of Anterior Nucleus of the Thalamus for Epilepsy Clinical Trial. Neurosurgery. 2021 Jun 23;89(3):406–12.

43. Schaper FLWVJ, Plantinga BR, Colon AJ, Wagner GL, Boon P, Blom N, et al. Deep Brain Stimulation in Epilepsy: A Role for Modulation of the Mammillothalamic Tract in Seizure Control? Neurosurgery. 2020 Sep;87(3):602.

44. Gompel JJV, Klassen BT, Worrell GA, Lee KH, Shin C, Zhao CZ, et al. Anterior nuclear deep brain stimulation guided by concordant hippocampal recording. 2015 Jun 1 [cited 2025 Apr 15]; Available from: https://thejns.org/focus/view/journals/neurosurg-focus/38/6/article-pE9.xml

45. Wang YC, Kremen V, Brinkmann BH, Middlebrooks EH, Lundstrom BN, Grewal SS, et al. Probing circuit of Papez with stimulation of anterior nucleus of the thalamus and hippocampal evoked potentials. Epilepsy Res. 2020 Jan;159:106248.

46. Aiello G, Ledergerber D, Dubcek T, Stieglitz L, Baumann C, Polanìa R, et al. Functional network dynamics between the anterior thalamus and the cortex in deep brain stimulation for epilepsy. Brain. 2023 Nov 1;146(11):4717–35.

47. Rolls ET. Chapter 2 - The cingulate cortex and limbic systems for action, emotion, and memory. In: Vogt BA, editor. Handbook of Clinical Neurology [Internet]. Elsevier; 2019 [cited 2024 Dec 17]. p. 23–37. (Cingulate Cortex; vol. 166). Available from: https://www.sciencedirect.com/science/article/pii/B9780444641960000029

48. Law N, Smith ML, Widjaja E. Thalamocortical Connections and Executive Function in Pediatric Temporal and Frontal Lobe Epilepsy. Am J Neuroradiol. 2018 Aug 1;39(8):1523–9.

49. Giampiccolo D, Binding LP, Caciagli L, Rodionov R, Foulon C, de Tisi J, et al. Thalamostriatal disconnection underpins long-term seizure freedom in frontal lobe epilepsy surgery. Brain. 2023 Jun 1;146(6):2377–88.

50. Tamura M, Spellman TJ, Rosen AM, Gogos JA, Gordon JA. Hippocampal-prefrontal theta-gamma coupling during performance of a spatial working memory task. Nat Commun. 2017 Dec 19;8(1):2182.

51. Sanger ZT, Zhang X, Leppik IE, Lisko T, Netoff TI, McGovern RA. Anterior nucleus of thalamus deep brain stimulation for medication refractory epilepsy modulates theta and low-frequency gamma activity: a case study. Ther Adv Neurol Disord. 2025 Jan 1;18:17562864251323052.

52. Johnson GW, Doss DJ, Morgan VL, Paulo DL, Cai LY, Shless JS, et al. The Interictal Suppression Hypothesis in focal epilepsy: network-level supporting evidence. Brain. 2023 Feb 1;146(7):2828–45.

53. Voges BR, Schmitt FC, Hamel W, House PM, Kluge C, Moll CKE, et al. Deep brain stimulation of anterior nucleus thalami disrupts sleep in epilepsy patients. Epilepsia. 2015 Aug;56(8):e99–103.

54. Swann NC, Hemptinne C de, Miocinovic S, Qasim S, Ostrem JL, Galifianakis NB, et al. Chronic multisite brain recordings from a totally implantable bidirectional neural interface: experience in 5 patients with Parkinson’s disease. 2017 Apr 14 [cited 2025 Feb 18]; Available from: https://thejns-org.ezp2.lib.umn.edu/view/journals/j-neurosurg/128/2/article-p605.xml

55. Rosin B, Slovik M, Mitelman R, Rivlin-Etzion M, Haber SN, Israel Z, et al. Closed-Loop Deep Brain Stimulation Is Superior in Ameliorating Parkinsonism. Neuron. 2011 Oct 20;72(2):370–84.

56. Stanslaski S, Herron J, Chouinard T, Bourget D, Isaacson B, Kremen V, et al. A Chronically-Implantable Neural Coprocessor for Investigating the Treatment of Neurological Disorders. IEEE Trans Biomed Circuits Syst. 2018 Dec;12(6):1230–45.

57. Sanger Z, Ventz S, McGovern R, Netoff T. Medtronic PerceptTM Recorded LFP Pre-Processing to Remove Noise and Cardiac Signals From Neural Recordings [Internet]. medRxiv; 2025 [cited 2025 Apr 14]. p. 2025.03.17.25324121. Available from: https://www.medrxiv.org/content/10.1101/2025.03.17.25324121v1

58. Donoghue T, Haller M, Peterson EJ, Varma P, Sebastian P, Gao R, et al. Parameterizing neural power spectra into periodic and aperiodic components. Nat Neurosci. 2020 Dec;23(12):1655–65.

59. Sanger Z. Neuromodulation for Refractory Epilepsy: Biomarkers and Stimulation Strategies [Internet] [Ph.D.]. [United States -- Minnesota]: University of Minnesota; 2025 [cited 2025 Aug 5]. Available from: https://www.proquest.com/docview/3225380657/abstract/85A1CBB846AC4584PQ/1

